# A National Health Service in ‘Serious Trouble’: what do multiple long-term conditions tell us about deterioration in health among people accessing hospital care in the North East of England?

**DOI:** 10.1101/2025.11.14.25340239

**Authors:** Rachel Cooper, Marzieh Shahmandi, Jonathan G Bunn, Peta Leroux, Chris Plummer, Tom P Marshall, James MS Wason, Miles D Witham, Avan A Sayer, the ADMISSION Research Collaborative

## Abstract

**Background:** The transformation of the National Health Service (NHS), outlined in the 10 Year Health Plan for England, must take account of the inexorable rise in the prevalence of multiple long-term conditions (MLTC). However, most evidence on MLTC is based on primary care data preceding the COVID-19 pandemic and, data from hospitals are scarce. We aimed to analyse routinely-collected electronic health records to estimate the prevalence of MLTC among adults accessing hospital care in North East England before and after the COVID-19 pandemic, and to test associations between sociodemographic characteristics and MLTC within these populations.

**Methods and Findings:** All adults with at least one admission to Newcastle upon Tyne Hospitals NHS Foundation Trust (NuTH) between July 2018 and June 2019 (N= 88,117, 51% women) or between July 2021 and June 2022 (N=83,036, 51% women) were included in analyses. MLTC was defined as the presence of two or more long-term conditions from an established list of 60 conditions. Poisson regression models were used to test associations of age, sex, ethnicity and index of multiple deprivation with MLTC.

Among people admitted to NuTH at least once between July 2018 and June 2019, overall prevalence of MLTC was 49.6% and between July 2021 and June 2022 it was 61.0%. Older age and neighbourhood deprivation were associated with increased risk of MLTC; people living in the most deprived neighbourhoods had a prevalence of MLTC equivalent to people from the least deprived neighbourhoods a decade older. Associations between neighbourhood deprivation and MLTC were stronger in younger adults; those aged 30-39y living in the most deprived neighbourhoods had 1.74 (95% confidence interval: 1.49-2.02) times higher risk of MLTC than those living in the least deprived neighbourhoods.

**Conclusions:** Among adults accessing inpatient hospital care in North East England, the prevalence of MLTC is high, has increased since the COVID-19 pandemic and has a greater impact on younger adults from the most deprived neighbourhoods. This highlights the sheer scale of the challenge that MLTC present in hospitals which must be addressed when redesigning services fit for the future.

## Introduction

In September 2024, Lord Darzi, presented the findings of his independent investigation of the National Health Service (NHS) in England^1^ to the Secretary of State for Health and Social Care. His key finding, that the ‘National Health Service is in serious trouble’ (p.1), is supported by evidence of substantial increases in waiting times^2^ and the lowest ever levels of patient satisfaction.^3,4^

The Darzi report (2024) highlighted a number of important failings and challenges which were already well recognised by people working in the NHS and those accessing its services. In doing this, the report shone a spotlight on some of the key factors that need to be addressed when transforming the NHS so that it once again functions efficiently and adequately meets the needs of the population that it was established to serve. It is imperative that this radical transformation, plans for which were outlined in the UK Government’s 10 year Health Plan for England launched in July 2025,^5 6^ is informed by robust empirical evidence.

Darzi’s report not only highlighted challenges within the NHS but also the important impact of the deterioration of the nation’s health on the NHS’ performance. Indicators of this deterioration include the stalling of long-term upward trends in life expectancy and declines in healthy life expectancy,^7 8^ which were exacerbated by but not fully explained by the COVID-19 pandemic. Of particular concern is that people living with disadvantage have experienced more rapid declines in these and other key indicators of population health and so inequalities are widening.^9^ It will therefore be essential to consider health inequalities and establish the NHS’s role in addressing these as part of its transformation.^9^

An important trend in population health underpinning declines in healthy life expectancy and further exacerbating health inequalities, is what Darzi refers to as a ‘surge in multiple long-term conditions’ (p.2). Multiple long-term conditions (MLTC), or multimorbidity as it is also known, is the co-occurrence of two or more chronic physical, mental or infectious health conditions.^10^ The inexorable rise in the prevalence of MLTC in recent decades, which data from primary care and population-based studies suggests has impacted people from more deprived neighbourhoods at younger ages,^11 12^ has been highlighted by the Chief Medical Officer for England, Professor Chris Whitty, as one of the greatest challenges facing medicine and science.^13–15^

It is widely recognised that transformation of the NHS must take account of the fact that many people in the population it serves are now living with two or more long-term conditions; analyses of primary care data estimated that one in four adults in England were living with MLTC in 2018^16^ and an analysis of 60 million GP records from 2020 reported prevalence estimates of MLTC ranging from 5.9% in 20-49 year olds to 68.2% in those aged 80 and over.^17^ The Darzi report presented NHS England population segmentation data showing an increase in prevalence of MLTC from 11.6% in 2017 to 15.5% in 2022^1^ and population projections suggest that the prevalence of MLTC will continue to rise.^18,19^ In this context MLTC in secondary care warrants special attention as radical transformation of hospital services will be required given their current configuration to deliver care for single conditions.^20^

Much of our current understanding of MLTC, and Darzi’s assertions on the scale of the challenge these present, is based on seminal studies of primary care data,^11,16,17,21^ on data preceding the COVID-19 pandemic,^22,23^ or from analyses focused on older adults.^24,25^ Evidence on MLTC in hospitals that is needed to inform transformation of the NHS is therefore limited. In analyses of 107 million admissions recorded in Hospital Episode Statistics between 2006 and 2021, prevalence of MLTC increased among both emergency and elective admissions in all age groups.^26^ These trends are indicative of the ‘nation’s deteriorating health’ but caution is required in interpreting the scale of the reported differences as individuals living with MLTC are at greater risk of hospital admission^27^ and so will have been counted multiple times in these analyses.

In analyses of primary care data from 2004 to 2019, the prevalence and incidence of MLTC were higher in the North East than in other regions of England.^12^ This aligns with evidence that the North East has poorer health, lower healthy life expectancy and greater inequalities than other regions in England.^28 29^ Addressing important gaps in our understanding of MLTC in hospitals by undertaking analyses within this region is therefore likely to maximise the impact of such research.

Using data on the index admissions of adults accessing planned and unplanned inpatient hospital care via the Newcastle upon Tyne Hospitals NHS Foundation Trust during two 12-month periods, before and after the COVID-19 pandemic, we aimed to estimate the prevalence of MLTC among the population accessing hospital care in the North East of England and examine differences in the distribution of MLTC by socio-demographic factors.

## Methods

Analyses were undertaken using routinely-collected data from electronic health records managed by the Newcastle upon Tyne Hospitals NHS Foundation Trust (NuTH), one of the busiest and largest NHS hospital trusts in England. The data are derived from records of all admissions to the Royal Victoria Infirmary and Freeman Hospital, hospitals based in Newcastle upon Tyne that provide a wide range of acute, general and specialist services for people living in urban and rural areas across the North East of England and beyond.

Working in close collaboration with the Digital Services team within NuTH, data were extracted on adult (18+ years) admissions for any reason, excluding obstetrics and regular day attenders, during two 12-month periods, 1 July 2018 to 30 June 2019 and 1 July 2021 to 30 June 2022. In both time periods the first admission for each individual was identified and for this index admission data were ascertained on the type of admission (classified as emergency, scheduled inpatient or day case), patient characteristics (age, sex, ethnicity and index of multiple deprivation) and all diagnoses, from a list of 60 long-term conditions (and their 2530 associated ICD-10 codes (see Supporting File S1)),^30^ coded and clinician recorded during the index admission spell or in lookback data covering any previous admissions to NuTH from 1^st^ January 2017 onwards.

### Ethics approval

Approval to access these data for research purposes was granted by the East Midlands-Derby Research Ethics Committee (20/EM/0186) and Confidentiality Advisory Group (21/CAG/0003) as part of umbrella governance covering all planned analyses within the ADMISSION Research Collaborative using routine healthcare data from NuTH. People who had chosen to opt out of the use or disclosure of their data for research and planning via the NHS National Data Opt-Out or locally were excluded. As data were de-identified, consent from individuals was not required.

### Defining multiple long-term conditions

In both time periods, patients were classified as living with MLTC if two or more of the 60 LTC were recorded in their electronic health record during the index admission or in lookback data. The process for identifying this list of 60 LTC is described elsewhere^30^ and was utilised to ensure a consistent and transparent approach to the definition and operationalisation of MLTC in the UK hospital setting.

### Covariates

Age in years was calculated using recorded date of birth and date of index admission. Recorded sex is assumed to reflect biological sex as self-reported gender was not routinely captured prior to 2024. Index of multiple deprivation (IMD) was derived from Post Code at the time of the index admission unless this information was missing because the patient had no fixed abode, did not report this or their main place of residence was outside England. Ethnicity was self-selected by patients from a list of 16 categories as per the national mandatory standard for the collection and analysis of ethnicity^31^ and recategorized for the purposes of analyses into five groups (White, South Asian, Black, Mixed, Other) (see Supporting Table S2).

### Patient and Public Involvement

Building on public engagement and priority setting work on long-term conditions led by the James Lind Alliance^32^ we have engaged with people with lived experience of MLTC throughout the lifetime of the ADMISSION Research Collaborative,^20^ which this study forms part of, benefiting from their input to all aspects of our work from funding bid development through to dissemination of findings. Adhering with UK Standards for Public Involvement, we established a Patient Advisory Group (PAG), including two public co-investigators and 14 other members of the public with lived experience of MLTC who were diverse in terms of age, gender, ethnicity and LTC, and met with them every four months (between early 2021 and summer 2025) and also consulted them on an ad hoc basis. During these regular interactions, we discussed their perspectives and sought advice on our research plans including the work presented in this paper.

### Statistical analyses

Once datasets had been compiled which included all relevant data from the index admission and available lookback files for all adults who had been admitted to NuTH at least once during two 12-month periods (July 1^st^ 2018 to June 30^th^ 2019 and July 1^st^ 2021 to June 30^th^ 2022, referred to as time periods 1 and 2), the sociodemographic characteristics of patients and prevalence of MLTC, counts of LTC and individual LTC in both time periods were summarised. Poisson regression models were then run to test the associations of age, sex, ethnicity and IMD with risk of MLTC in both time periods, unadjusted and adjusted for all other socio-demographic characteristics. Formal tests of interaction between age and IMD were undertaken in fully-adjusted models and stratified results presented where appropriate.

Data from the two time periods were then pooled, allowing formal tests of interaction between time period and each socio-demographic characteristic to be undertaken and for the association between time period and MLTC to be estimated with adjustment for socio-demographic characteristics.

As maximum lookback time for diagnoses was greater in time period 2 than time period 1, sensitivity analyses were undertaken in which descriptive analyses were rerun for time period 2 with restriction of lookback time to the same maximum length as time period 1.

All analyses were conducted in R (version 4.4.3; R Core Team, 2025) using the tidyverse package for data wrangling and manipulation, ggplot2 for visualisation, and lmtest for comparing nested Poisson models using likelihood ratio tests. All analysis code is publicly available on GitHub at https://github.com/shahmandi/MLTC_Admission_Descriptive_Article (commit ID: 68e2e029d95e0a96dcd9ec7b470d1b99b0938248) and has been fully annotated for ease of use and reproducibility.

## Results

During July 2018 to June 2019 and July 2021 to June 2022, 88,117 and 83,036 people, respectively, aged 18 years and over were admitted at least once to NuTH. During both time periods just over 50% of patients were women, mean age was 59 years, most were White and almost one third lived in the most deprived neighbourhoods (table 1). Between July 2018 and June 2019, 32.7% of patients’ index admissions were classified as an emergency, and this increased to 40.9% among patients admitted between July 2021 and June 2022. The distribution of characteristics was similar in men and women in both time periods (Supporting Table S3).

**Table 1:** Characteristics of people with at least one recorded admission to Newcastle upon Tyne Hospitals NHS Foundation Trust during two 12-month periods (01/07/2018 - 30/06/2019 and 01/07/2021 - 30/06/2022)

|  |  | N (%) |  |
| --- | --- | --- | --- |
|  |  | Time period 1<br>2018 – 2019<br>(N=88,117) | Time period 2<br>2021 – 2022<br>(N=83,036) |
| **Sex** |  |  |  |
|  | Men | 42,791 (48.6) | 40,846 (49.2) |
|  | Women | 45,317 (51.4) | 42,171 (50.8) |
|  | Missing | 9 | 19 |
| **Mean age (y) (SD)** |  | 58.5 (19.2) | 58.9 (19.5) |
| **Age group (y)** |  |  |  |
|  | 18 - 29 | 9021 (10.2) | 8301 (10.0) |
|  | 30 – 39 | 8817 (10.0) | 8495 (10.2) |
|  | 40 – 49 | 9278 (10.5) | 8614 (10.4) |
|  | 50 – 59 | 14738 (16.7) | 12912 (15.6) |
|  | 60 – 69 | 16551 (18.8) | 15563 (18.8) |
|  | 70 – 79 | 17019 (19.3) | 16665 (20.1) |
|  | 80 – 89 | 10676 (12.1) | 10101 (12.2) |
|  | 90+ | 1984 (2.3) | 2269 (2.7) |
|  | Missing | 33 | 116 |
| **Ethnicity** |  |  |  |
|  | White | 76645 (95.1) | 70,833 (94.3) |
|  | South Asian | 1844 (2.3) | 1907 (2.5) |
|  | Black | 456 (0.6) | 567 (0.8) |
|  | Mixed | 319 (0.4) | 320 (0.4) |
|  | Other | 1304 (1.6) | 1428 (1.9) |
|  | Missing | 7549 | 7981 |
| **Index of multiple deprivation** |  |  |  |
|  | 1-2 (most deprived) | 28186 (32.1) | 26426 (32.0) |
|  | 3-4 | 18005 (20.5) | 16691 (20.2) |
|  | 5-6 | 13942 (15.9) | 13523 (16.4) |
|  | 7-8 | 12683 (14.5) | 12041 (14.6) |
|  | 9-10 (least deprived) | 14915 (17.0) | 14042 (17.0) |
|  | Missing | 386 | 313 |
| **Index admission type** |  |  |  |
|  | Emergency | 28,805 (32.7) | 33,943 (40.9) |
|  | Scheduled inpatient | 17,390 (19.7) | 14,729 (17.7) |
|  | Day case | 41,922 (47.6) | 34,364 (41.4) |

See Supporting Table S3 for equivalent statistics stratified by sex

Among people admitted to NuTH at least once between July 2018 and June 2019, overall prevalence of MLTC was 49.6% and between July 2021 and June 2022 it was 61.0% (table 2). Prevalence of 4 or more LTC increased from 20% to 32%. When stratified by age group, prevalence of MLTC in time period 2 ranged from 21% in people aged 18-29 years to 92% in those aged 90 years and over (see Supporting Table S4). For prevalence estimates stratified by sex see Supporting Table S5 and for prevalence of MLTC by type of index admission see Supporting Table S6.

**Table 2:** Count of long-term conditions (LTC) and prevalence of multiple long-term conditions (MLTC) among people with at least one recorded admission to Newcastle upon Tyne Hospitals NHS Foundation Trust during two 12-month periods (01/07/2018 - 30/06/2019 and 01/07/2021 - 30/06/2022)

|  |  | N (%) |  |
| --- | --- | --- | --- |
|  |  | Time period 1<sup>†</sup><br>2018 – 2019<br>(N=88,117) | Time period 2<sup>‡</sup><br>2021 – 2022<br>(N=83,036) |
| **No. of LTC\*** | 0 | 23968 (27.2) | 17299 (20.8) |
|  | 1 | 20418 (23.2) | 15087 (18.2) |
|  | 2 | 15093 (17.1) | 13221 (15.9) |
|  | 3 | 11345 (12.9) | 10839 (13.1) |
|  | 4 | 7475 (8.5) | 8444 (10.2) |
|  | 5 | 4499 (5.1) | 6083 (7.3) |
|  | 6 | 2592 (2.9) | 4288 (5.2) |
|  | 7 | 1430 (1.6) | 2841 (3.4) |
|  | 8 | 704 (0.8) | 1950 (2.4) |
|  | 9+ | 593 (0.7) | 2984 (3.6) |
| **MLTC** |  |  |  |
| 0-1 LTC |  | 44,386 (50.4) | 32,386 (39.0) |
| ≥ 2 LTC |  | 43,731 (49.6) | 50,650 (61.0) |

\* Total number of conditions diagnosed and coded in each person's electronic health record (during the index admission or lookback) from a potential list of 60 LTC<sup>30</sup>
† Lookback time range (y): 0 – 1.4; mean (SD): 0.5 (0.3)
‡ Lookback time range (y): 0 – 4.4; mean (SD): 2.4 (1.1)

Between 2018-2019 and 2021-2022 increases were observed in the prevalence of 54 of the 60 LTC used to define MLTC (table 3). In both time periods, the most prevalent LTC was hypertension. Eight other LTC (coronary artery disease, diabetes mellitus, solid organ cancers, arrhythmia, osteoarthritis, asthma, depression and chronic kidney disease) were ranked in the top 10 in both time periods, with chronic obstructive pulmonary disease also in the top 10 in time period 1 replaced by anxiety in time period 2.

**Table 3:**
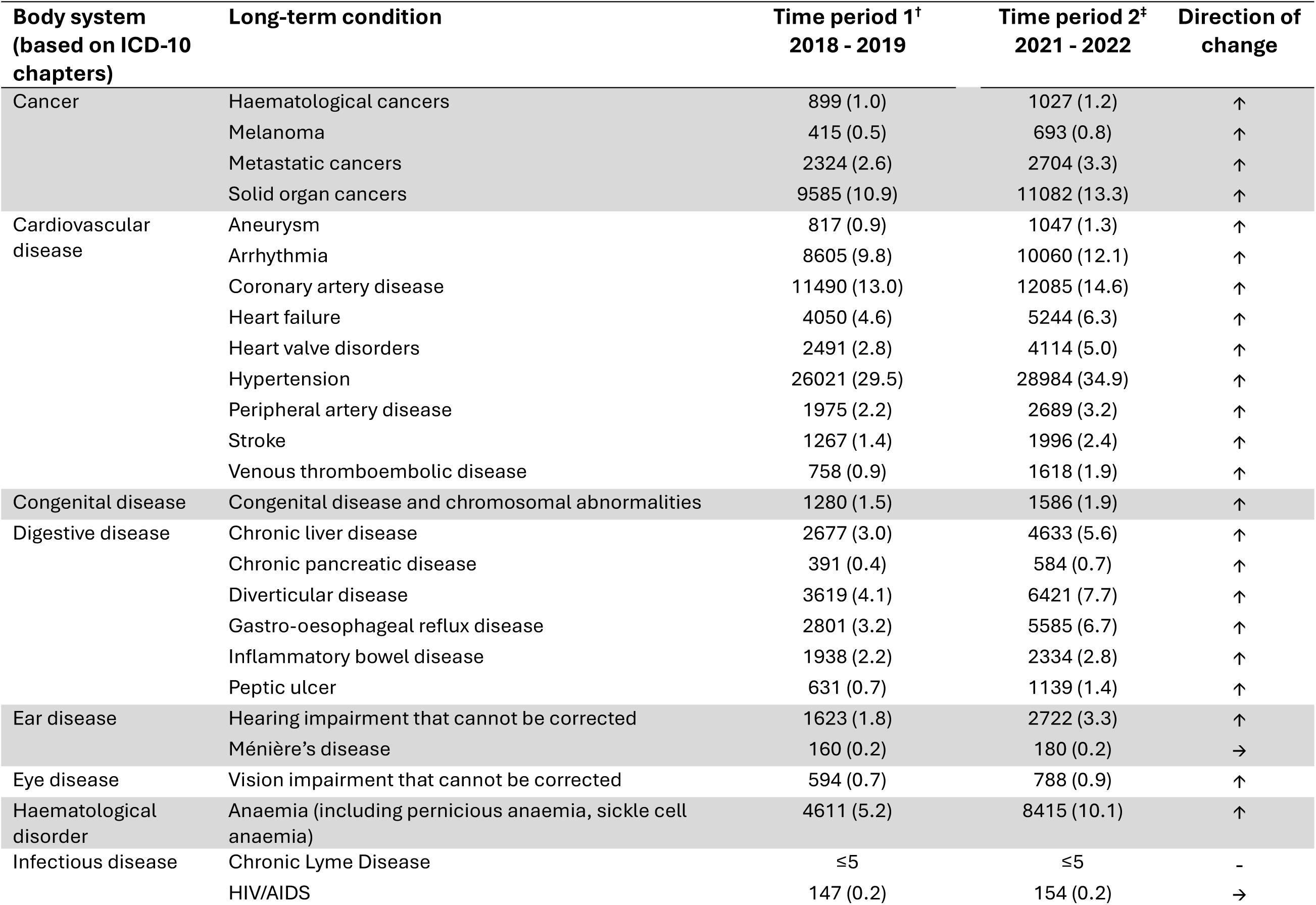
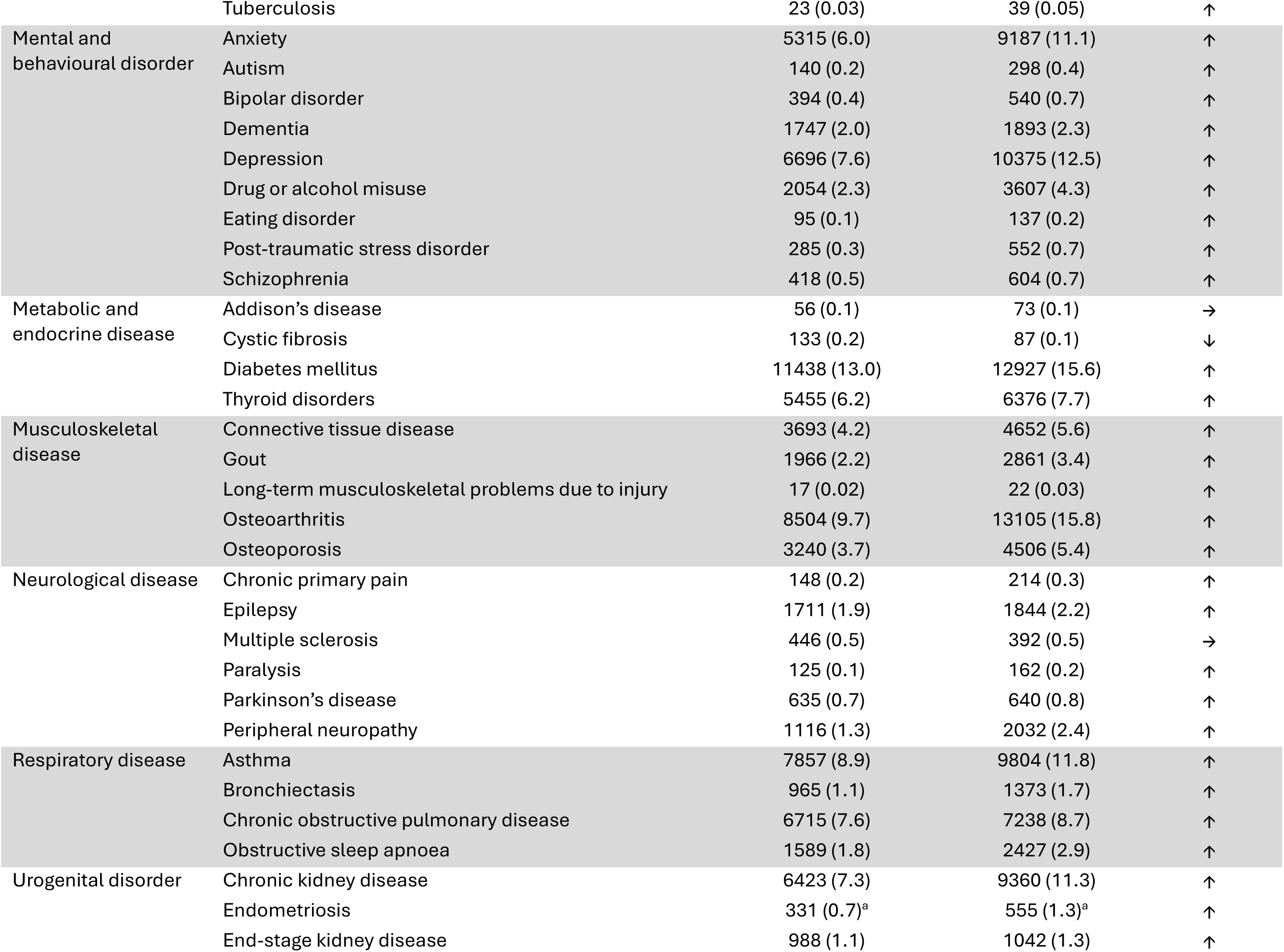
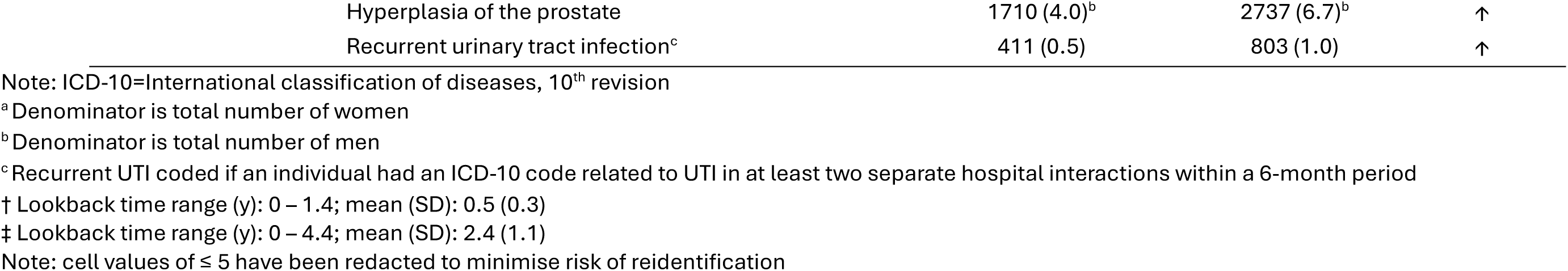
Prevalence of 60 long-term conditions among people with at least one recorded admission to Newcastle upon Tyne Hospitals NHS Foundation Trust during two 12-month periods (01/07/2018 - 30/06/2019 (N=88,117) and 01/07/2021 - 30/06/2022 (N=83,036))

In time period 1, women had lower risk of MLTC than men but there was no association between sex and MLTC in time period 2 (p<0.001 for test of interaction between sex and time period) (table 4). In both time periods there was a clear and consistent association between older age and increased risk of MLTC. Associations between ethnicity and MLTC observed in unadjusted models, which showed lower risk of MLTC in most minoritised ethnic groups when compared with the White ethnic group, were largely explained by adjustment for age, sex and IMD (table 4). In models adjusted for sex, age and ethnicity, there was evidence of increased risk of MLTC among people living in more deprived neighbourhoods. However, overall estimates need to be interpreted with caution due to evidence of interaction between age and IMD (p<0.001 for tests of interaction in both time periods). When models of the association between IMD and MLTC were stratified by age group, there was consistent evidence in both time periods of stronger associations between IMD and MLTC in younger age groups (figure 1 and Supporting Figure S7 and Supporting Table S8); for example among adults aged 30-39 years admitted to hospital in either time period, those living in the most deprived neighbourhoods had 1.74 times higher risk of MLTC when compared with those living in the least deprived neighbourhoods (95% Confidence Intervals (CI): 1.45 to 2.08 and 1.49 to 2.02, for time periods 1 and 2, respectively). Much smaller associations between IMD and MLTC were observed in people aged 70 to 89 and no associations were observed in those aged 90 and over.

**Table 4:** Associations between socio-demographic characteristics and risk of MLTC in time periods 1 and 2 estimated using Poisson regression among samples with complete data on all covariates

|  |  | Risk ratio of MLTC (95% CI) |  |  |  |
| --- | --- | --- | --- | --- | --- |
|  |  | Time period 1<br>2018 – 2019<br>(N=80,171) |  | Time period 2<br>2021 - 2022<br>(N=74,656) |  |
|  |  | Unadjusted | Mutually adjusted | Unadjusted | Mutually adjusted |
| **Sex** | Men | 1.00 | 1.00 | 1.00 | 1.00 |
|  | Women | 0.92 (0.90, 0.94) | 0.95 (0.93, 0.97) | 0.96 (0.95, 0.98) | 0.99 (0.98, 1.01) |
| **Age group (y)** | 18 - 29 | 1.00 | 1.00 | 1.00 | 1.00 |
|  | 30 – 39 | 1.43 (1.33, 1.55) | 1.43 (1.33, 1.55) | 1.44 (1.35, 1.54) | 1.43 (1.35, 1.53) |
|  | 40 – 49 | 2.28 (2.13, 2.44) | 2.29 (2.14, 2.45) | 2.05 (1.93, 2.17) | 2.04 (1.93, 2.17) |
|  | 50 – 59 | 3.09 (2.91, 3.30) | 3.09 (1.90, 3.29) | 2.67 (2.53, 2.82) | 2.65 (2.51, 2.80) |
|  | 60 – 69 | 4.21 (3.96, 4.48) | 4.24 (3.99, 4.51) | 3.25 (3.08, 3.42) | 3.24 (3.08, 3.42) |
|  | 70 – 79 | 4.99 (4.70, 5.31) | 5.06 (4.77, 5.38) | 3.69 (3.50, 3.88) | 3.71 (3.52, 3.91) |
|  | 80 – 89 | 5.55 (5.22, 5.91) | 5.65 (5.31, 6.01) | 4.03 (3.83, 4.25) | 4.07 (3.86, 4.30) |
|  | 90+ | 5.92 (5.49, 6.39) | 6.09 (5.64, 6.57) | 4.21 (3.94, 4.50) | 4.27 (4.00, 4.56) |
| **Index of multiple deprivation†** | 9-10 (least deprived) | 1.00 | 1.00 | 1.00 | 1.00 |
|  | 7-8 | 1.04 (1.00, 1.08) | 1.07 (1.03, 1.11) | 1.00 (0.97, 1.04) | 1.04 (1.00, 1.07) |
|  | 5-6 | 1.03 (1.00, 1.07) | 1.11 (1.07, 1.15) | 1.00 (0.97, 1.04) | 1.07 (1.04, 1.10) |
|  | 3-4 | 1.06 (1.02, 1.09) | 1.18 (1.14, 1.22) | 1.01 (0.98, 1.04) | 1.12 (1.09, 1.15) |
|  | 1-2 (most deprived) | 1.11 (1.08, 1.14) | 1.29 (1.26, 1.33) | 1.07 (1.04, 1.10) | 1.23 (1.19, 1.26) |
| **Ethnicity** | White | 1.00 | 1.00 | 1.00 | 1.00 |
|  | South Asian | 0.76 (0.70, 0.81) | 0.96 (0.89, 1.03) | 0.77 (0.73, 0.83) | 0.94 (0.88, 1.01) |
|  | Black | 0.51 (0.43, 0.61) | 0.81 (0.68, 0.97) | 0.59 (0.52, 0.68) | 0.80 (0.70, 0.92) |
|  | Mixed | 0.57 (0.46, 0.70) | 0.90 (0.73, 1.11) | 0.60 (0.51, 0.72) | 0.85 (0.71, 1.02) |
|  | Other | 0.53 (0.48, 0.59) | 0.75 (0.68, 0.84) | 0.57 (0.52, 0.62) | 0.76 (0.70, 0.83) |

Note: analyses restricted to samples with complete data on age, sex, ethnicity and IMD; mutually adjusted models include adjustment for all other covariates
† Interactions between age and IMD $p < 0.001$ - please see Figure 1 and Supporting files S7 and S8 for associations between IMD and MLTC stratified by age group

In models of the association between time period and MLTC, the unadjusted relative risk of MLTC in time period 2 (July 2021-June 2022) when compared with time period 1 (July 2018-June 2019) was 1.24 (95% CI: 1.22 to 1.25). After adjustment for age, sex, IMD and ethnicity this association was maintained (RR: 1.23 (95% CI: 1.21 to 1.24)).

In sensitivity analyses, prevalence estimates of MLTC and individual LTC in time period 2 were slightly reduced when lookback time was restricted to a maximum of 1.4 years (the maximum lookback for time period 1) (e.g. overall prevalence of MLTC was 58% and prevalence of 4 or more LTC was 28%) but were still higher than equivalent estimates for time period 1 (Supporting Tables S9 and S10).

## Discussion

Among adults accessing hospital care in the North East of England, the prevalence of MLTC is high and has increased markedly since the COVID-19 pandemic. Increases have been observed in a wide range of physical and mental LTC across different body systems in adults of all ages.

Between July 2021 and June 2022 almost two thirds of people admitted to NuTH at least once had two or more LTC and almost a third had four or more. In younger adults, prevalence of MLTC was above 20% and by 60-69 it had reached 70%, with figures even higher among those adults in each age group living in the most deprived neighbourhoods. These estimates have been produced using a standard list of 60 LTC tailored for use in a hospital setting^30^ but are still likely to be conservative given evidence that identifying diagnoses for LTC from hospital record data alone can lead to underestimates in their prevalence.^33^

These findings demonstrate how the trend of deteriorating population health related to increasing prevalence of MLTC^1,34^ is manifesting in a hospital setting. In addition, the findings highlight the importance of recognising and addressing the sheer scale of the challenge that MLTC present in hospitals when implementing the UK Government’s 10 year Health Plan for England.^5^

Published prevalence estimates of MLTC vary markedly, due to differences in population characteristics and methods.^35,36^ However, a comparison of our prevalence estimates with those published in 2024 using national primary care data for England up to March 2020, reporting a range in prevalence of MLTC from 5.9% in adults aged 20-49 to 68.2% in those aged 80 and over, highlights the value in generating robust evidence on MLTC in different settings; in planning for the transformation of the NHS, in particular hospital services, we risk underestimating the scale of the challenge that MLTC pose if we focus on estimates from community-dwelling populations and primary care data which have until now been more widely used to characterise MLTC in the UK than data from hospitals.^36,37^

The increase in prevalence of MLTC observed over a relatively short time frame, spanning the COVID-19 pandemic, highlights the importance of characterising MLTC among people accessing health care post-COVID, as we have done, for future planning. While overall patterns of associations between socio-demographic characteristics and MLTC did not change markedly, those people who accessed hospital care after the peak of the COVID-19 pandemic had accumulated more LTC than those accessing hospital care shortly before, a trend also reflected in analyses of Hospital Episode Statistics.^26^

Our study highlights the power of harnessing information at a local level, something which will be increasingly required if the Government is to deliver on its ambition of a new operating model for the NHS that redistributes ‘power from the centre to the frontline’.^5^ By focusing on data from a specific hospital trust in an area of the country with a high level of need and, working in collaboration with the hospital’s coding and digital services teams, we avoid the limitation of differences in coding practices between hospitals.^26^ In addition we can have confidence that the increase in prevalence we have observed is unlikely to be explained by changes in diagnosis, recording or coding practices. That increases were observed for the majority of LTC supports this notion, as any nationally mandated changes in coding practice would have been expected to impact some LTC more than others. In our main analyses, we utilised the maximum lookback time available for each time period with the aim of providing the most comprehensive estimates of MLTC prevalence. As there was longer lookback for time period 2, caution is required in interpreting the absolute scale of the difference in prevalence of MLTC between the two time periods, due to greater potential for informed presence bias in the latter. However, sensitivity analyses showed a higher prevalence of MLTC in time period 2 than in time period 1 even when the maximum lookback time was the same.

Differences in MLTC prevalence between the two time periods were not explained by adjustment for sociodemographic characteristics. This suggests differences in the prevalence of MLTC between time periods 1 and 2 are unlikely to be due to changes in the socio-demographic structure of the patient population or minor differences in the strength of the associations between socio-demographic characteristics and MLTC between time periods. A higher proportion of index admissions were classified as emergency in time period 2 than 1, a trend which has been consistently observed post-COVID. Although prevalence of MLTC varied by type of index admission, increases in prevalence of MLTC over time were observed for all three types of index admission (Supporting Table S6) suggesting that increased use of emergency hospital services is unlikely to explain our findings. Our aim in this paper was to undertake an overall assessment of MLTC prevalence among people accessing hospital care for any reason and so we focused on the first admission of individuals admitted to hospital at least once for any reason within a specified time period. However, the differences in prevalence of MLTC among people accessing different types of hospital services we observe warrants further investigation in future analyses focusing on specific types of hospital admission (rather than first admission of any type). Exploring different combinations and patterns of LTC to aid identification of the most impactful MLTC in populations accessing hospital care also requires further research given our finding that MLTC, defined as 2 or more LTC, is ubiquitous in this setting, especially at older ages.

In the UK Government’s 10 year Health Plan for England launched in July 2025,^5^ three major shifts were outlined. Our findings have implications for all three. In accelerating the first shift - from analogue to digital - it will be essential to ensure that MLTC is captured and characterised consistently in electronic health records. Our work demonstrates that this is possible using data within a hospital trust. The next step will be to ensure that this is incorporated into electronic health records in real-time so that it can inform clinical decision making and empower patients, especially where this could facilitate the coordination and integration of care that people living with MLTC admitted to hospital have consistently highlighted as lacking.^38^ In planning for the second shift - from hospital to community - our findings highlight the importance of considering the high prevalence of MLTC in the population currently accessing hospital care, especially those from more deprived neighbourhoods in regions with poorer health and greater inequalities. A comprehensive understanding of the scale of the challenge MLTC presents, and of the needs of people currently accessing hospital care, is required to realise the ambition of providing more of the health care people require in their own communities and to ensure resources allocated to achieve this are adequately aligned with level of need within regions. Finally, the evidence we present supports the third shift - from sickness to prevention. As outlined by Fraser and Alwan,^39^ prevention of MLTC needs to be prioritised. As a large and increasing proportion of people are living with MLTC, we need to prevent incident cases of MLTC and also mitigate the impacts of MLTC for those people already living with MLTC with a clear focus on ensuring interventions are equitable. This will be essential to ease current and future pressures on the NHS and address health inequity.

**Figure 1:**
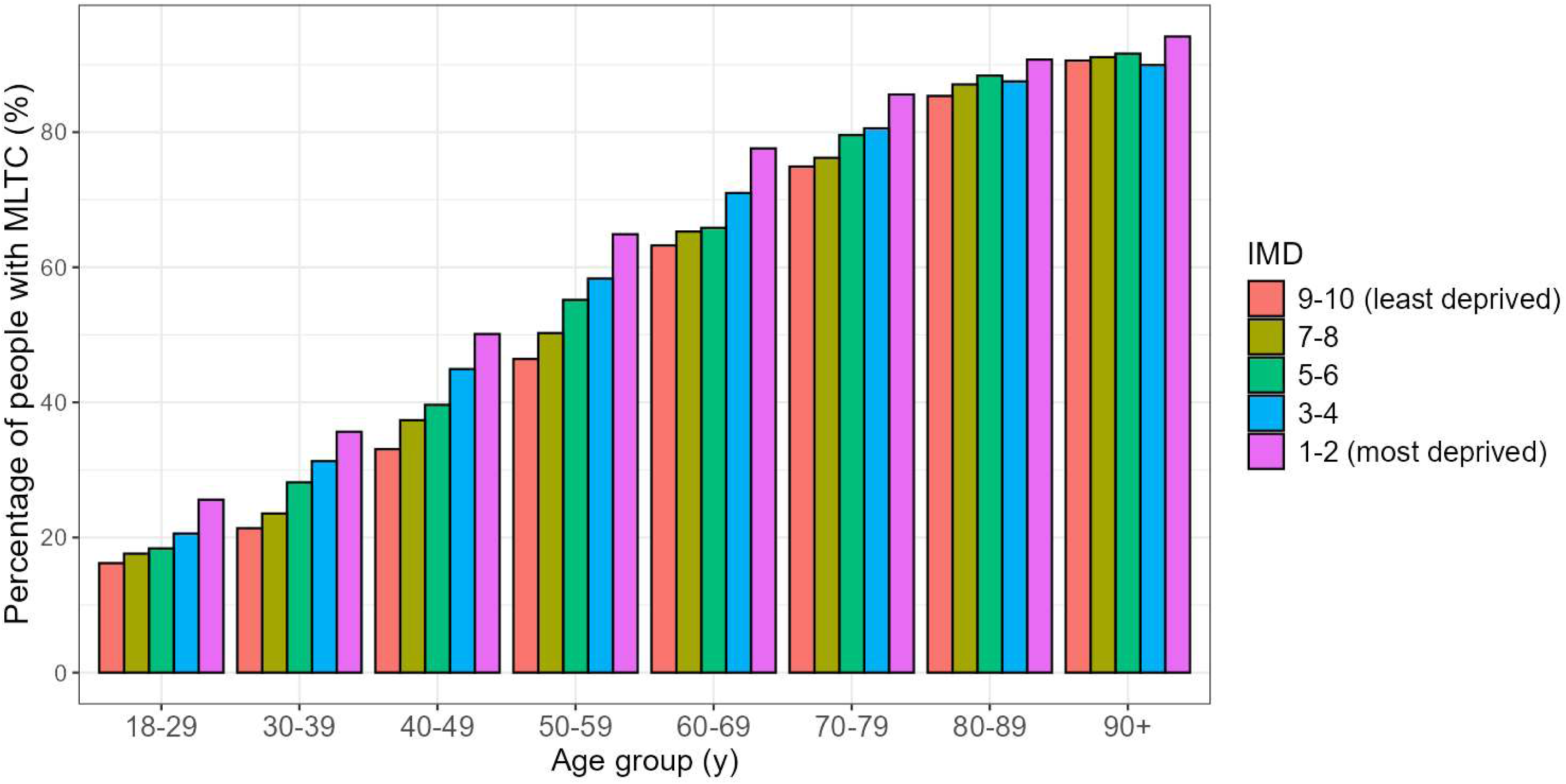
Prevalence of MLTC by age and index of multiple deprivation among people with at least one recorded admission to Newcastle upon Tyne Hospitals NHS Foundation Trust during time period 2 (01/07/2021 - 30/06/2022) with complete data on all covariates (N=74,656) Footnote: Please see Supporting Figure S7 for equivalent figure for time period 1 and Supporting Table S8 for results from Poisson regression models of the associations between IMD and risk of MLTC stratified by age group in both time periods.

## Data Availability

All data produced in the present work are contained in the manuscript

## Acknowledgements

This work uses data provided by patients and collected by the NHS as part of their care and support.

The authors thank Hayley Richardson and Allison McCall from the Newcastle upon Tyne Hospitals NHS Foundation Trust clinical coding team for reviewing our code lists.

## Authors’ contributions

AAS (PI) with RC, MDW, CP, TPM and JMSW (co-Is) acquired the funding for this study. RC, MDW and AAS conceived the idea for the study and along with MS, JGB, TPM and JMSW developed the study objectives. PL and CP prepared the data, oversaw access and provided guidance on its use. MS and JGB undertook quality checks on the data and prepared a dataset for analyses. MS undertook the main analyses, supervised by RC and JMSW. RC prepared the first draft of the manuscript. All authors critically reviewed and edited the manuscript and approved the final version for submission.

RC is responsible for the overall content as guarantor and accepts full responsibility for the work and/or the conduct of the study, had access to the data, and controlled the decision to publish. The corresponding author attests that all listed authors meet authorship criteria and that no others meeting the criteria have been omitted.

