## Supplementary files 2 to 10 for "A National Health Service in ‘Serious Trouble’: what do multiple long-term conditions tell us about deterioration in health among people accessing hospital care in the North East of England?"

**Table S2:** Five high-level ethnicity groups used in analyses and their component categories

| High-level ethnicity categories used in analyses | Component categories |
|---|---|
| White | White–British<br>White–Irish<br>White–Any other white background |
| South Asian | Asian or Asian British-Indian<br>Asian or Asian British-Pakistani<br>Asian or Asian British–Bangladeshi<br>Asian or Asian British–Any other Asian background |
| Black | Black or Black British-Caribbean<br>Black or Black British–African<br>Black or Black British–Any other Black background |
| Mixed | Mixed–White and Black Caribbean<br>Mixed–White and Black African<br>Mixed–White and Asian<br>Mixed–Any other mixed background |
| Other | Other Ethnic groups–Chinese<br>Other Ethnic groups–Any other ethnic group |

Component categories self-selected by patients from a standard list of 16 options[1] and grouped for the purposes of analyses to maintain statistical power, ensuring alignment with previous analyses of electronic health records[2]

**Table S3:** Characteristics of people with at least one recorded admission to Newcastle upon Tyne Hospitals NHS Foundation Trust during two 12-month periods (01/07/2018 - 30/06/2019 and 01/07/2021 - 30/06/2022) stratified by sex

| | N (%) | | | |
|---|---|---|---|---|
| | **Time period 1 2018 – 2019** | | **Time period 2 2021 - 2022** | |
| | **Men (N=42,791)** | **Women (N=45,317)** | **Men (N=40,846)** | **Women (N=42,171)** |
| **Mean age (y) (SD)** | 59.0 (18.7) | 58.0 (19.7) | 59.5 (18.9) | 58.3 (20.0) |
| **Age group (y)** | | | | |
| 18 - 29 | 4102 (9.6) | 4916 (10.9) | 3767 (9.2) | 4528 (10.8) |
| 30 – 39 | 3865 (9.0) | 4951 (10.9) | 3753 (9.2) | 4740 (11.3) |
| 40 – 49 | 4295 (10.0) | 4981 (11.0) | 4022 (9.9) | 4589 (10.9) |
| 50 – 59 | 7310 (17.1) | 7427 (16.4) | 6461 (15.8) | 6449 (15.3) |
| 60 – 69 | 8728 (20.4) | 7821 (17.3) | 8324 (20.4) | 7235 (17.2) |
| 70 – 79 | 8698 (20.3) | 8321 (18.4) | 8784 (21.5) | 7880 (18.7) |
| 80 – 89 | 5014 (11.7) | 5662 (12.5) | 4785 (11.7) | 5315 (12.6) |
| 90+ | 763 (1.8) | 1221 (2.7) | 881 (2.2) | 1388 (3.3) |
| *Missing* | *16* | *17* | *69* | *47* |
| **Ethnicity** | | | | |
| White | 36952 (95.2) | 39686 (95.1) | 34629 (94.7) | 36198 (94.1) |
| South Asian | 846 (2.2) | 998 (2.4) | 828 (2.3) | 1077 (2.8) |
| Black | 197 (0.5) | 259 (0.6) | 231 (0.6) | 336 (0.9) |
| Mixed | 149 (0.4) | 169 (0.4) | 149 (0.4) | 170 (0.4) |
| Other | 682 (1.8) | 622 (1.5) | 737 (2.0) | 690 (1.8) |
| *Missing* | *3965* | *3583* | *4272* | *3700* |
| **Index of multiple deprivation** | | | | |
| 1-2 (most deprived) | 13409 (31.5) | 14773 (32.7) | 12723 (31.3) | 13700 (32.6) |
| 3-4 | 8786 (20.6) | 9217 (20.4) | 8263 (20.3) | 8423 (20.0) |
| 5-6 | 6839 (16.1) | 7103 (15.7) | 6727 (16.6) | 6792 (16.2) |
| 7-8 | 6301 (14.8) | 6381 (14.1) | 6008 (14/8) | 6030 (14.3) |
| 9-10 (least deprived) | 7225 (17.0) | 7688 (17.0) | 6924 (17.0) | 7117 (16.9) |
| *Missing* | *231* | *155* | *201* | *109* |
| **Index admission type** | | | | |
| Emergency | 14460 (33.8) | 14340 (31.6) | 16921 (41.4) | 17011 (40.3) |
| Scheduled inpatient | 8939 (20.9) | 8450 (18.7) | 7671 (18.8) | 7056 (16.7) |
| Day case | 19392 (45.3) | 22527 (49.7) | 16254 (39.8) | 18104 (42.9) |

**Table S4:** Prevalence of multiple long-term conditions (MLTC) among people with at least one recorded admission to Newcastle upon Tyne Hospitals NHS Foundation Trust during two 12-month periods (01/07/2018 - 30/06/2019 and 01/07/2021 - 30/06/2022) stratified by age group

| | Prevalence of MLTC - N (%) | |
|---|---|---|
| | **Time period 1† 2018 – 2019** | **Time period 2‡ 2021 – 2022** |
| **Age group (y)** | | |
| 18 - 29 | 1253 (13.9) | 1748 (21.1) |
| 30 – 39 | 1737 (19.7) | 2599 (30.6) |
| 40 – 49 | 2934 (31.6) | 3758 (43.6) |
| 50 – 59 | 6291 (42.7) | 7391 (57.2) |
| 60 – 69 | 9703 (58.6) | 10896 (70.0) |
| 70 – 79 | 11852 (69.6) | 13300 (79.8) |
| 80 – 89 | 8315 (77.9) | 8878 (87.9) |
| 90+ | 1646 (83.0) | 2079 (91.6) |

† Lookback time range (y): 0 – 1.4; mean (SD): 0.5 (0.3)
‡ Lookback time range (y): 0 – 4.4; mean (SD): 2.4 (1.1)

**Table S5:** Count of long-term conditions (LTC) and prevalence of multiple long-term conditions (MLTC) among people with at least one recorded admission to Newcastle upon Tyne Hospitals NHS Foundation Trust during two 12-month periods (01/07/2018 - 30/06/2019 and 01/07/2021 - 30/06/2022) stratified by sex

| | N (%) | | | |
|---|---|---|---|---|
| | Time period 1† 2018 – 2019 | | Time period 2‡ 2021 - 2022 | |
| | Men (N=42,791) | Women (N=45,317) | Men (N=40,846) | Women (N=42,171) |
| **No. of LTC*** | | | | |
| 0 | 11093 (25.9) | 12870 (28.4) | 8422 (20.6) | 8870 (21.0) |
| 1 | 9625 (22.5) | 10792 (23.8) | 7108 (17.4) | 7972 (18.9) |
| 2 | 7416 (17.3) | 7676 (16.9) | 6337 (15.5) | 6884 (16.3) |
| 3 | 5729 (13.4) | 5615 (12.4) | 5408 (13.2) | 5430 (12.9) |
| 4 | 3816 (8.9) | 3659 (8.1) | 4298 (10.5) | 4145 (9.8) |
| 5 | 2370 (5.5) | 2129 (4.7) | 3070 (7.5) | 3013 (7.1) |
| 6 | 1355 (3.2) | 1236 (2.7) | 2201 (5.4) | 2087 (5.0) |
| 7 | 744 (1.7) | 686 (1.5) | 1512 (3.7) | 1327 (3.2) |
| 8 | 333 (0.8) | 371 (0.8) | 989 (2.4) | 961 (2.3) |
| 9+ | 310 (0.7) | 283 (0.6) | 1501 (3.7) | 1482 (3.5) |
| **MLTC** | | | | |
| 0-1 LTC | 20718 (48.4) | 23662 (52.2) | 15530 (38.0) | 16842 (39.9) |
| ≥ 2 LTC | 22073 (51.6) | 21655 (47.8) | 25316 (62.0) | 25329 (60.1) |

* Total number of conditions diagnosed and coded in each person's electronic health record (during the index admission or lookback) from a potential list of 60 LTC[3]

† Lookback time range (y): 0 – 1.4; mean (SD): 0.5 (0.3)

‡ Lookback time range (y): 0 – 4.4; mean (SD): 2.4 (1.1)

**Table S6:** Prevalence of MLTC and characteristics of people with at least one recorded admission to Newcastle upon Tyne Hospitals NHS Foundation Trust during two 12-month periods (01/07/2018 - 30/06/2019 and 01/07/2021 - 30/06/2022) stratified by type of index admission

| | N (%) | | | | | |
|---|---|---|---|---|---|---|
| | Time period 1† 2018 – 2019 | | | Time period 2‡ 2021 – 2022 | | |
| **Type of index admission** | **Emergency** | **Scheduled inpatient** | **Day case** | **Emergency** | **Scheduled inpatient** | **Day Case** |
| **Total N** | 28805 | 17390 | 41922 | 33943 | 14729 | 34364 |
| **Sex** | | | | | | |
| Men | 14460 (50.2) | 8939 (51.4) | 19392 (46.3) | 16921 (49.9) | 7671 (52.1) | 16254 (47.3) |
| Women | 14340 (49.8) | 8450 (48.6) | 22527 (53.7) | 17011 (50.1) | 7056 (47.9) | 18104 (52.7) |
| *Missing* | *5* | *1* | *3* | *11* | *2* | *6* |
| **Mean age (y) (SD)** | 56.6 (21.7) | 59.4 (17.1) | 59.4 (18.1) | 56.2 (21.7) | 60.4 (17.1) | 60.8 (17.8) |
| **Ethnicity** | | | | | | |
| White | 25499 (93.4) | 15107 (97.1) | 36039 (95.6) | 28910 (92.1) | 12512 (96.8) | 29411 (95.6) |
| South Asian | 799 (2.9) | 185 (1.2) | 860 (2.3) | 1045 (3.3) | 185 (1.4) | 677 (2.2) |
| Black | 217 (0.8) | 67 (0.4) | 172 (0.5) | 358 (1.1) | 63 (0.5) | 146 (0.5) |
| Mixed | 126 (0.5) | 51 (0.3) | 142 (0.4) | 178 (0.6) | 34 (0.3) | 108 (0.4) |
| Other | 665 (2.4) | 142 (0.9) | 497 (1.3) | 887 (2.8) | 127 (1.0) | 414 (1.4) |
| *Missing* | *1499* | *1838* | *4212* | *2565* | *1808* | *3608* |
| **IMD** | | | | | | |
| 1-2 (most deprived) | 11351 (39.7) | 5000 (28.9) | 11835 (28.3) | 12659 (37.5) | 4096 (27.9) | 9671 (28.2) |
| 3-4 | 5493 (19.2) | 3876 (22.4) | 8636 (20.7) | 6301 (18.7) | 3369 (23.0) | 7021 (20.5) |
| 5-6 | 4179 (14.6) | 2996 (17.3) | 6767 (16.2) | 5222 (15.5) | 2605 (17.8) | 5696 (16.6) |
| 7-8 | 3462 (12.1) | 2614 (15.1) | 6607 (15.8) | 4396 (13.0) | 2262 (15.4) | 5383 (15.7) |
| 9-10 (least deprived) | 4086 (14.3) | 2846 (16.4) | 7983 (19.1) | 5171 (15.3) | 2337 (15.9) | 6534 (19.1) |
| *Missing* | *234* | *58* | *94* | *194* | *60* | *59* |
| **MLTC** | | | | | | |
| 0-1 LTC | 13843 (48.1) | 6503 (37.4) | 24040 (57.3) | 13628 (40.2) | 4309 (29.3) | 14449 (42.1) |
| ≥ 2 LTC | 14962 (51.9) | 10887 (62.6) | 17882 (42.7) | 20315 (59.9) | 10420 (70.7) | 19915 (58.0) |

† Lookback time range (y): 0 – 1.4; mean (SD): 0.5 (0.3) ‡ Lookback time range (y): 0 – 4.4; mean (SD): 2.4 (1.1)

**Figure S7:** Prevalence of MLTC by age and index of multiple deprivation among people with at least one recorded admission to Newcastle upon Tyne Hospitals NHS Foundation Trust during time period 1 (01/07/2018 - 30/06/2019) with complete data on all covariates (N=80,171)

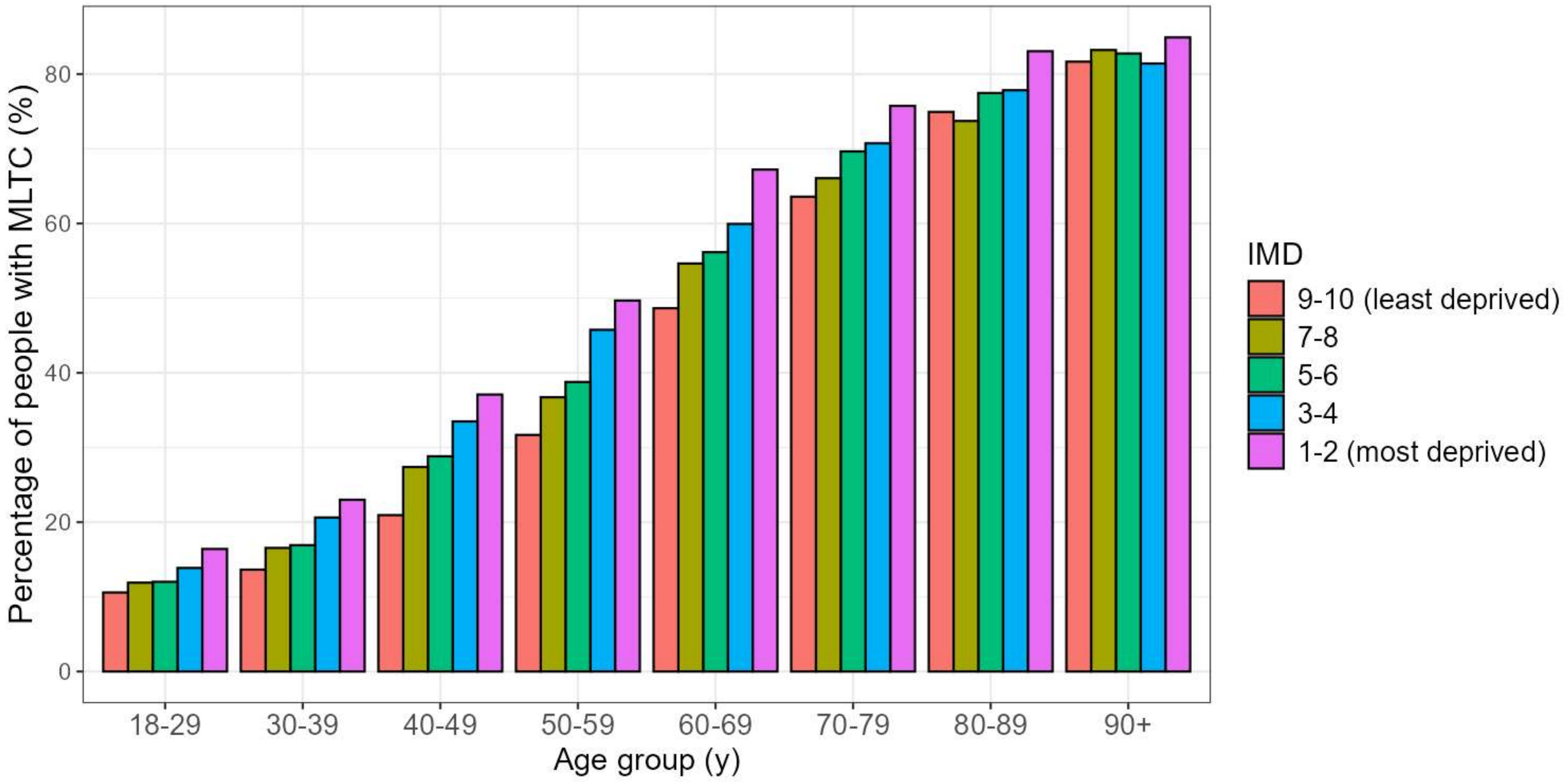

**Table S8:** Associations between index of multiple deprivation and risk of MLTC stratified by age group in time periods 1 and 2 estimated using Poisson regression among samples with complete data on all covariates

| | Adjusted* Risk Ratio of MLTC (95% CI) | | | | | | | |
|---|---|---|---|---|---|---|---|---|
| **Age group (y)** | **18-29** | **30-39** | **40-49** | **50-59** | **60-69** | **70-79** | **80-89** | **90+** |
| **Time period 1 (2018-2019) (N=80,171)** | | | | | | | | |
| 9-10 (least deprived) | 1.00 | 1.00 | 1.00 | 1.00 | 1.00 | 1.00 | 1.00 | 1.00 |
| 7-8 | 1.15 (0.88, 1.50) | 1.23 (0.98, 1.55) | 1.32 (1.11, 1.56) | 1.15 (1.03, 1.28) | 1.13 (1.04, 1.22) | 1.04 (0.98, 1.11) | 0.98 (0.91, 1.05) | 1.02 (0.87, 1.19) |
| 5-6 | 1.17 (0.91, 1.51) | 1.24 (1.00, 1.54) | 1.41 (1.21, 1.66) | 1.21 (1.10, 1.35) | 1.15 (1.06, 1.23) | 1.10 (1.04, 1.17) | 1.03 (0.96, 1.11) | 1.02 (0.86, 1.20) |
| 3-4 | 1.36 (1.08, 1.72) | 1.55 (1.27, 1.88) | 1.62 (1.40, 1.87) | 1.44 (1.31, 1.58) | 1.22 (1.14, 1.31) | 1.12 (1.05, 1.19) | 1.03 (0.96, 1.11) | 0.99 (0.85, 1.16) |
| 1-2 (most deprived) | 1.62 (1.31, 2.01) | 1.74 (1.45, 2.08) | 1.83 (1.59, 2.09) | 1.55 (1.43, 1.69) | 1.36 (1.28, 1.45) | 1.19 (1.12, 1.25) | 1.10 (1.04, 1.18) | 1.03 (0.90, 1.19) |
| **Time period 2 (2021-2022) (N=74,656)** | | | | | | | | |
| 9-10 (least deprived) | 1.00 | 1.00 | 1.00 | 1.00 | 1.00 | 1.00 | 1.00 | 1.00 |
| 7-8 | 1.13 (0.91, 1.40) | 1.14 (0.94, 1.39) | 1.11 (0.96, 1.29) | 1.08 (0.98, 1.19) | 1.04 (0.97, 1.12) | 1.02 (0.96, 1.08) | 1.02 (0.95, 1.09) | 1.01 (0.88, 1.16) |
| 5-6 | 1.19 (0.96, 1.46) | 1.38 (1.16, 1.65) | 1.19 (1.04, 1.36) | 1.19 (1.08, 1.30) | 1.05 (0.97, 1.12) | 1.05 (0.99, 1.11) | 1.03 (0.96, 1.10) | 1.01 (0.88, 1.16) |
| 3-4 | 1.34 (1.11, 1.62) | 1.49 (1.26, 1.76) | 1.34 (1.18, 1.52) | 1.26 (1.16, 1.37) | 1.12 (1.05, 1.20) | 1.08 (1.02, 1.14) | 1.02 (0.95, 1.09) | 1.00 (0.86, 1.15) |
| 1-2 (most deprived) | 1.63 (1.36, 1.94) | 1.74 (1.49, 2.02) | 1.54 (1.37, 1.72) | 1.39 (1.29, 1.50) | 1.23 (1.16, 1.31) | 1.14 (1.08, 1.20) | 1.06 (0.99, 1.12) | 1.04 (0.92, 1.17) |

* Adjusted for sex and ethnicity; p-values for formal tests of interaction between IMD and age <0.001 in both time periods

**Table S9:** Count of long-term conditions (LTC) and prevalence of multiple long-term conditions (MLTC) among people with at least one recorded admission to Newcastle upon Tyne Hospitals NHS Foundation Trust during time period 2 (01/07/2021 - 30/06/2022), with lookback time for ascertainment of diagnoses restricted to a maximum of 1.4 years

| | **N (%)**<br>**Time period 2†**<br>**2021 – 2022**<br>**(N=83.036)** |
|---|---|
| **No. of LTC*** | |
| 0 | 18933 (22.8) |
| 1 | 16336 (19.7) |
| 2 | 13756 (16.6) |
| 3 | 11063 (13.3) |
| 4 | 8239 (9.9) |
| 5 | 5684 (6.9) |
| 6 | 3676 (4.4) |
| 7 | 2249 (2.7) |
| 8 | 1418 (1.7) |
| 9+ | 1682 (2.0) |
| **MLTC** | |
| 0-1 LTC | 35,269 (42.5) |
| ≥ 2 LTC | 47,767 (57.5) |

* Total number of conditions diagnosed and coded in each person's electronic health record (during the index admission or lookback) from a potential list of 60 LTC[3]
† Lookback time restricted to a maximum of 1.4 years for comparison with maximum lookback time available for people admitted during time period 1

**Table S10:** Prevalence of 60 long-term conditions among people with at least one recorded admission to Newcastle upon Tyne Hospitals NHS Foundation Trust during time period 2 (01/07/2021 - 30/06/2022 (N=83,036)), with lookback time for ascertainment of diagnoses restricted to a maximum of 1.4 years

| **Body system (based on ICD-10 chapters)** | **Long-term condition** | **Time period 2 (restricted lookback) 2021 - 2022** |
|---|---|---|
| Cancer | Haematological cancers | 914 (1.1) |
| | Melanoma | 538 (0.6) |
| | Metastatic cancers | 2451 (3.0) |
| | Solid organ cancers | 9116 (11.0) |
| Cardiovascular disease | Aneurysm | 925 (1.1) |
| | Arrhythmia | 9503 (11.4) |
| | Coronary artery disease | 11295 (13.6) |
| | Heart failure | 4555 (5.5) |
| | Heart valve disorders | 3600 (4.3) |
| | Hypertension | 27185 (32.7) |
| | Peripheral artery disease | 2375 (2.9) |
| | Stroke | 1571 (1.9) |
| | Venous thromboembolic disease | 1216 (1.5) |
| Congenital disease | Congenital disease and chromosomal abnormalities | 1366 (1.6) |
| Digestive disease | Chronic liver disease | 4097 (4.9) |
| | Chronic pancreatic disease | 498 (0.6) |
| | Diverticular disease | 5335 (6.4) |
| | Gastro-oesophageal reflux disease | 4415 (5.3) |
| | Inflammatory bowel disease | 2200 (2.6) |
| | Peptic ulcer | 745 (0.9) |
| Ear disease | Hearing impairment that cannot be corrected | 2227 (2.7) |
| | Ménière's disease | 147 (0.2) |
| Eye disease | Vision impairment that cannot be corrected | 648 (0.8) |
| Haematological disorder | Anaemia (including pernicious anaemia, sickle cell anaemia) | 6848 (8.2) |
| Infectious disease | Chronic Lyme Disease | ≤5 |
| | HIV/AIDS | 149 (0.2) |
| | Tuberculosis | 30 (0.04) |
| Mental and behavioural disorder | Anxiety | 7758 (9.3) |
| | Autism | 278 (0.3) |
| | Bipolar disorder | 486 (0.6) |
| | Dementia | 1831 (2.2) |
| | Depression | 8767 (10.6) |
| | Drug or alcohol misuse | 3183 (3.8) |
| | Eating disorder | 108 (0.1) |
| | Post-traumatic stress disorder | 468 (0.6) |
| | Schizophrenia | 541 (0.7) |
| Metabolic and | Addison's disease | 64 (0.1) |

| | | |
|---|---|---|
| endocrine disease | Cystic fibrosis | 85 (0.1) |
| | Diabetes mellitus | 12574 (15.1) |
| | Thyroid disorders | 5922 (7.1) |
| Musculoskeletal disease | Connective tissue disease | 4270 (5.1) |
| | Gout | 2413 (2.9) |
| | Long-term musculoskeletal problems due to injury | 17 (0.02) |
| | Osteoarthritis | 11236 (13.5) |
| | Osteoporosis | 3953 (4.8) |
| Neurological disease | Chronic primary pain | 174 (0.2) |
| | Epilepsy | 1701 (2.0) |
| | Multiple sclerosis | 385 (0.5) |
| | Paralysis | 138 (0.2) |
| | Parkinson's disease | 625 (0.8) |
| | Peripheral neuropathy | 1585 (1.9) |
| Respiratory disease | Asthma | 8813 (10.6) |
| | Bronchiectasis | 1209 (1.5) |
| | Chronic obstructive pulmonary disease | 6758 (8.1) |
| | Obstructive sleep apnoea | 2165 (2.6) |
| Urogenital disorder | Chronic kidney disease | 8698 (10.5) |
| | Endometriosis | 439 (1.0)[a] |
| | End-stage kidney disease | 1014 (1.2) |
| | Hyperplasia of the prostate | 2223 (5.4)[b] |
| | Recurrent urinary tract infection[c] | 571 (0.7) |

Note: ICD-10=International classification of diseases, 10th revision

[a] Denominator is total number of women

[b] Denominator is total number of men

[c] Recurrent UTI coded if an individual had an ICD-10 code related to UTI in at least two separate hospital interactions within a 6-month period

Note: cell values of ≤ 5 have been redacted to minimise risk of reidentification
